# Machine learning-based prediction of cognitive outcomes in de novo Parkinson’s disease

**DOI:** 10.1101/2022.02.02.22270300

**Authors:** Joshua Harvey, Rick A Reijnders, Rachel Cavill, Annelien Duits, Sebastian Köhler, Lars Eijssen, Bart PF Rutten, Gemma Shireby, Ali Torkamani, Byron Creese, Albert FG Leentjens, Katie Lunnon, Ehsan Pishva

**Author notes:** Correspondence to: Ehsan Pishva, Department of Psychiatry and Neuropsychology, School for Mental Health and Neuroscience, Maastricht University, P.O. Box 616, 6200 MD Maastricht, The Netherlands. These authors contributed equally to this work (Joshua Harvey, Rick A Reijnders).

## Abstract

Cognitive impairment is a debilitating symptom in Parkinson’s disease (PD). We aimed to establish an accurate multivariate machine learning (ML) model to predict cognitive outcome in newly diagnosed PD cases from the Parkinson’s Progression Markers Initiative (PPMI). Annual cognitive assessments over an eight-year time span were used to define two cognitive outcomes of i) cognitive impairment, and ii) dementia conversion. Selected baseline variables were organized into three subsets of clinical, biofluid and genetic/epigenetic measures and tested using four different ML algorithms. Irrespective of the ML algorithm used, the models consisting of the clinical variables performed best and showed better prediction of cognitive impairment outcome over dementia conversion. We observed a marginal improvement in the prediction performance when clinical, biofluid, and epigenetic/genetic variables were all included in one model. Several cerebrospinal fluid measures and an epigenetic marker showed high predictive weighting in multiple models when included alongside clinical variables.

## Introduction

Cognitive impairment and dementia are highly common and debilitating non-motor symptoms in Parkinson’s Disease (PD). Cognitive impairment in PD carries distinct diagnostic challenges, a higher burden of care, worse functioning, and a lower quality of life^1^. Cross-sectional population studies show that approximately 30% of cases with PD have dementia, with 20-25% of patients presenting with mild cognitive impairment (MCI)^2^ as early as diagnosis^3^. Longitudinal studies report an average of 50% of PD patients develop dementia within 10 years^4,5^. Several demographic and clinical measures have been shown to be predictive in PD cognitive impairment, including age, visual hallucinations, REM sleep disorder and severity of parkinsonism, in particular non-tremor symptoms^1^. Moreover, considerable research interest has focused on identifying objective biomarkers, including structural and functional imaging, biofluid measures and genetic risk^6-8^.

Recently, approaches for biomarker development have focused on the use of multivariate prediction, utilizing algorithms that combine multiple measures for individual-level cognitive outcome prediction^9,10^. Using multivariate panels of data, however, comes with challenges implicit in the complexity of multi-modal data. A growing area of research utilizes machine learning (ML) approaches both to identify data-driven subtypes of disease^11,12^ and to predict disease progression^13-15^ including future cognitive outcomes^10,16^.

A major challenge for predicting cognitive outcome in PD is the high levels of heterogeneity implicit within the condition, with high inter-individual variation in clinical presentation and progression^17^. In the present study, we assessed longitudinal records of cognitive diagnoses in the Parkinson’s Progression Markers Initiative (PPMI)^18^, a well-characterized cohort of early PD patients and used multiple ML methods to predict cognitive outcome using baseline variables. We assessed prediction of two outcome measures over an eight-year time period: i) development of cognitive impairment (MCI or dementia) and ii) development of dementia. Variables were split into three subsets, including clinical measures, biofluid (CSF, serum) assays and variables of genetic/epigenetic markers in blood.

For prediction, we applied four different machine learning algorithms (Random Forest [RF], ElasticNet, Support Vector Machines [SVM] and Conditional inference forest [Cforest]) and assessed the performance of each to determine if different learning approaches show better overall predictive accuracy. Applying multiple outcome measures, different subsets of predicting variables and ML algorithms, we aimed to test which showed the best overall predictive performance, establish powerful multivariate predictive models, and highlight important predictive variables included in these models.

## Results

### Prediction of cognitive outcomes

Using records of cognitive diagnosis over an eight-year time period (Figure 1), we subset two cognitive outcomes. The first outcome tested development of overall cognitive impairment, including a group showing solely normal or subjective cognitive decline (SCD) (n = 127) and another with development of MCI and Dementia (n= 82). The second outcome tested dementia development; comparing a dementia conversion group (n = 43) to a set of combined normal, SCD and MCI cases (n = 166) (Figure 1). Four ML algorithms were used for prediction using baseline variables, with each evaluated based on metrics of overall accuracy. Descriptive statistical summaries of each cognitive outcome group tested are shown in Table 1. Baseline variables were binned into individual subsets of genetic/epigenetic (47 variables), biofluid (12 variables) and clinical (64 variables) measures (Summarized in Table S1) and tested individually and collectively. An overview of individual ML algorithm accuracy for each variable subset and outcome are summarized in Figure 2 and Table 2.

**Table 1.** Summary statistics of demographic and selected clinical measures. For each outcome, summary values of mean for continuous measurements or proportions for categorical variables. Significance values reported as the results of a Mann-Whitney U test for continuous and a Chi-2 test for categorical variables (* P < 0.5, ** P < 1.0 E-3, *** P< 1.0 E-5). PD: Parkinson’s Disease, MDS-UPDRS: Movement Disorder Society Unified Parkinson’s Disease Rating Scale, HVLT: Hopkins Verbal Learning Test, STAI: State Trait Anxiety Inventory.

|  | Cognitive Impairment |  | Dementia Conversion |  |
| --- | --- | --- | --- | --- |
| Variable Name | Cognitively Intact | Cognitively Impaired | Non-Dementia | Dementia |
| Age at Baseline | 60.0(9.14) | 66.4(8.68) *** | 61.6(9.58) | 66.4(8.03) |
| Gender (Female/Male) | 45/82 | 18/66* | 53/114 | 10/34 |
| Years of Education | 15.9(2.76) | 15.6(3.17) | 16.0(2.69) | 15.0(3.64) |
| Duration of Disease since Diagnosis (Months) | 6.21(6.43) | 7.47(7.14) | 6.90(6.66) | 6.01(7.05) ** |
| Age at PD Diagnosis | 59.5(9.13) | 65.8(8.75) *** | 61.0(9.55) | 65.9(8.13) ** |
| Hoehn & Yahr Stage (0/1/2/3) | 0/67/59/1 | 0/33/51/0 | 0/82/84/1 | 0/18/26/0 |
| MDS-UPDRS Part III Score (OFF) | 18.8(7.8) | 22.7(8.9) * | 19.7(8.23) | 22.7(8.92) ** |
| Benton Judgement of Line Orientation Score | 13.4(1.64) | 12.0(2.47) ** | 13.1(1.96) | 11.8(2.41) *** |
| Geriatric Depression Scale Score | 1.91(2.23) | 2.90(2.45) ** | 2.22(2.42) | 2.61(2.18) |
| HVLT Immediate/Total Recall | 26.7(4.40) | 20.9(5.03) *** | 25.3(5.29) | 20.8(4.67) *** |
| HVLT Delayed Recall | 9.48(1.90) | 6.74(2.83) *** | 8.79(2.53) | 6.86(2.69) *** |
| HVLT Delayed Recognition | 11.5(0.789) | 10.6(1.510) *** | 11.2(1.170) | 10.8(1.360) ** |
| HVLT False Alarms | 0.976(1.02) | 1.520(1.38) ** | 1.050(1.12) | 1.730(1.39) ** |
| HVLT Discrimination Recognition | 10.40(1.59) | 8.69(2.84) *** | 10.00(2.1) | 8.59(2.81) *** |
| HVLT Retention | 0.913(0.132) | 0.786(0.278) ** | 0.881(0.192) | 0.789(0.267) ** |
| Letter Number Sequencing Score | 11.20(2.56) | 9.04(2.59) *** | 10.80(2.68) | 8.80(2.66) *** |
| Semantic Fluency Total Score | 51.9(10.60) | 41.3(9.12) *** | 49.6(11.10) | 40.3(8.96) *** |
| STAI Total Score | 61.6(15.5) | 70.2(18.1) ** | 63.7(16.1) | 70.0(19.7) ** |
| Symbol Digit Modalities Score | 44.6(7.43) | 34.2(9.64) *** | 42.7(8.49) | 31.8(9.63) *** |
| MOCA Score (adjusted for education) | 27.9(1.74) | 26.0(2.82) *** | 27.3(2.23) | 26.4(2.93) ** |

**Table 2.**
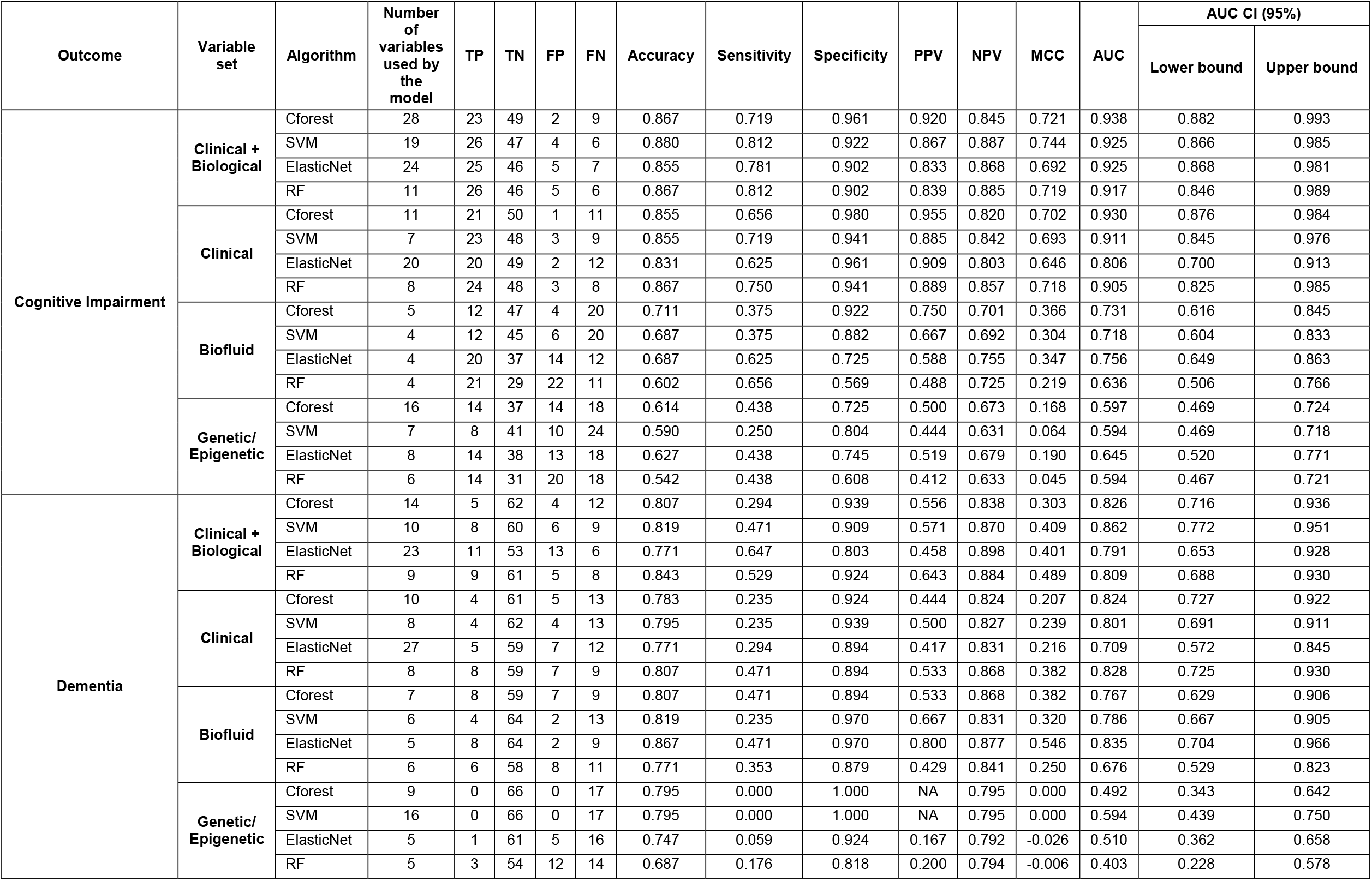
Summary of predictive accuracy for individual ML methods. Summary table of metrics evaluating accuracy of ML predictions. Abbreviation: TP: TruePositive (impaired/dementia), TN: True Negative (cognitively intact/non dementia), FP: False Positive, FN: False Negative. PPV: Positive Predictive Value, NPV: Negative Predictive Value. Lower and higher confidence intervals (CI) show 95% CI for AUC.

| Outcome | Variable set | Algorithm | Number of variables used by the model | TP | TN | FP | FN | Accuracy | Sensitivity | Specificity | PPV | NPV | MCC | AUC | AUC CI (95%) |  |
| --- | --- | --- | --- | --- | --- | --- | --- | --- | --- | --- | --- | --- | --- | --- | --- | --- |
|  |  |  |  |  |  |  |  |  |  |  |  |  |  |  | Lower bound | Upper bound |
| Cognitive Impairment | Clinical + Biological | Cforest | 28 | 23 | 49 | 2 | 9 | 0.867 | 0.719 | 0.961 | 0.920 | 0.845 | 0.721 | 0.938 | 0.882 | 0.993 |
|  |  | SVM | 19 | 26 | 47 | 4 | 6 | 0.880 | 0.812 | 0.922 | 0.867 | 0.887 | 0.744 | 0.925 | 0.866 | 0.985 |
|  |  | ElasticNet | 24 | 25 | 46 | 5 | 7 | 0.855 | 0.781 | 0.902 | 0.833 | 0.868 | 0.692 | 0.925 | 0.868 | 0.981 |
|  |  | RF | 11 | 26 | 46 | 5 | 6 | 0.867 | 0.812 | 0.902 | 0.839 | 0.885 | 0.719 | 0.917 | 0.846 | 0.989 |
|  | Clinical | Cforest | 11 | 21 | 50 | 1 | 11 | 0.855 | 0.656 | 0.980 | 0.955 | 0.820 | 0.702 | 0.930 | 0.876 | 0.984 |
|  |  | SVM | 7 | 23 | 48 | 3 | 9 | 0.855 | 0.719 | 0.941 | 0.885 | 0.842 | 0.693 | 0.911 | 0.845 | 0.976 |
|  |  | ElasticNet | 20 | 20 | 49 | 2 | 12 | 0.831 | 0.625 | 0.961 | 0.909 | 0.803 | 0.646 | 0.806 | 0.700 | 0.913 |
|  |  | RF | 8 | 24 | 48 | 3 | 8 | 0.867 | 0.750 | 0.941 | 0.889 | 0.857 | 0.718 | 0.905 | 0.825 | 0.985 |
|  | Biofluid | Cforest | 5 | 12 | 47 | 4 | 20 | 0.711 | 0.375 | 0.922 | 0.750 | 0.701 | 0.366 | 0.731 | 0.616 | 0.845 |
|  |  | SVM | 4 | 12 | 45 | 6 | 20 | 0.687 | 0.375 | 0.882 | 0.667 | 0.692 | 0.304 | 0.718 | 0.604 | 0.833 |
|  |  | ElasticNet | 4 | 20 | 37 | 14 | 12 | 0.687 | 0.625 | 0.725 | 0.588 | 0.755 | 0.347 | 0.756 | 0.649 | 0.863 |
|  |  | RF | 4 | 21 | 29 | 22 | 11 | 0.602 | 0.656 | 0.569 | 0.488 | 0.725 | 0.219 | 0.636 | 0.506 | 0.766 |
|  | Genetic/ Epigenetic | Cforest | 16 | 14 | 37 | 14 | 18 | 0.614 | 0.438 | 0.725 | 0.500 | 0.673 | 0.168 | 0.597 | 0.469 | 0.724 |
|  |  | SVM | 7 | 8 | 41 | 10 | 24 | 0.590 | 0.250 | 0.804 | 0.444 | 0.631 | 0.064 | 0.594 | 0.469 | 0.718 |
|  |  | ElasticNet | 8 | 14 | 38 | 13 | 18 | 0.627 | 0.438 | 0.745 | 0.519 | 0.679 | 0.190 | 0.645 | 0.520 | 0.771 |
|  |  | RF | 6 | 14 | 31 | 20 | 18 | 0.542 | 0.438 | 0.608 | 0.412 | 0.633 | 0.045 | 0.594 | 0.467 | 0.721 |
| Dementia | Clinical + Biological | Cforest | 14 | 5 | 62 | 4 | 12 | 0.807 | 0.294 | 0.939 | 0.556 | 0.838 | 0.303 | 0.826 | 0.716 | 0.936 |
|  |  | SVM | 10 | 8 | 60 | 6 | 9 | 0.819 | 0.471 | 0.909 | 0.571 | 0.870 | 0.409 | 0.862 | 0.772 | 0.951 |
|  |  | ElasticNet | 23 | 11 | 53 | 13 | 6 | 0.771 | 0.647 | 0.803 | 0.458 | 0.898 | 0.401 | 0.791 | 0.653 | 0.928 |
|  |  | RF | 9 | 9 | 61 | 5 | 8 | 0.843 | 0.529 | 0.924 | 0.643 | 0.884 | 0.489 | 0.809 | 0.688 | 0.930 |
|  | Clinical | Cforest | 10 | 4 | 61 | 5 | 13 | 0.783 | 0.235 | 0.924 | 0.444 | 0.824 | 0.207 | 0.824 | 0.727 | 0.922 |
|  |  | SVM | 8 | 4 | 62 | 4 | 13 | 0.795 | 0.235 | 0.939 | 0.500 | 0.827 | 0.239 | 0.801 | 0.691 | 0.911 |
|  |  | ElasticNet | 27 | 5 | 59 | 7 | 12 | 0.771 | 0.294 | 0.894 | 0.417 | 0.831 | 0.216 | 0.709 | 0.572 | 0.845 |
|  |  | RF | 8 | 8 | 59 | 7 | 9 | 0.807 | 0.471 | 0.894 | 0.533 | 0.868 | 0.382 | 0.828 | 0.725 | 0.930 |
|  | Biofluid | Cforest | 7 | 8 | 59 | 7 | 9 | 0.807 | 0.471 | 0.894 | 0.533 | 0.868 | 0.382 | 0.767 | 0.629 | 0.906 |
|  |  | SVM | 6 | 4 | 64 | 2 | 13 | 0.819 | 0.235 | 0.970 | 0.667 | 0.831 | 0.320 | 0.786 | 0.667 | 0.905 |
|  |  | ElasticNet | 5 | 8 | 64 | 2 | 9 | 0.867 | 0.471 | 0.970 | 0.800 | 0.877 | 0.546 | 0.835 | 0.704 | 0.966 |
|  |  | RF | 6 | 6 | 58 | 8 | 11 | 0.771 | 0.353 | 0.879 | 0.429 | 0.841 | 0.250 | 0.676 | 0.529 | 0.823 |
|  | Genetic/ Epigenetic | Cforest | 9 | 0 | 66 | 0 | 17 | 0.795 | 0.000 | 1.000 | NA | 0.795 | 0.000 | 0.492 | 0.343 | 0.642 |
|  |  | SVM | 16 | 0 | 66 | 0 | 17 | 0.795 | 0.000 | 1.000 | NA | 0.795 | 0.000 | 0.594 | 0.439 | 0.750 |
|  |  | ElasticNet | 5 | 1 | 61 | 5 | 16 | 0.747 | 0.059 | 0.924 | 0.167 | 0.792 | -0.026 | 0.510 | 0.362 | 0.658 |
|  |  | RF | 5 | 3 | 54 | 12 | 14 | 0.687 | 0.176 | 0.818 | 0.200 | 0.794 | -0.006 | 0.403 | 0.228 | 0.578 |

**Figure 1.**
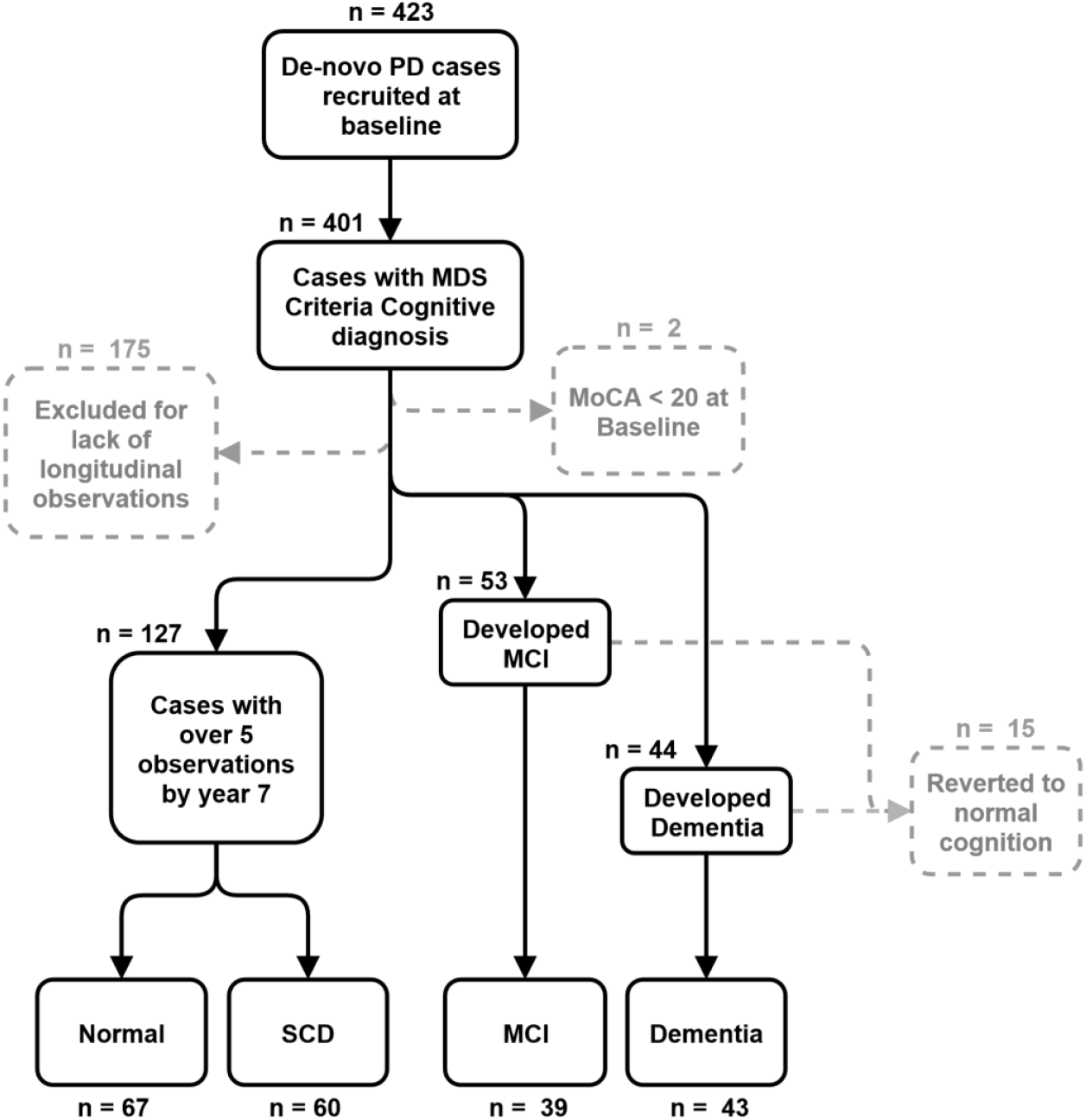
Flow diagram of case sub-setting criteria: Samples retained in each stage are shown as black lines between boxes, samples excluded shown as dotted grey lines and boxes. Case numbers for each selection stage are shown overlaid on each plot. Final subset groups (Normal, SCD, MCI and Dementia) are shown at the bottom of the flow diagram.

**Figure 2.**
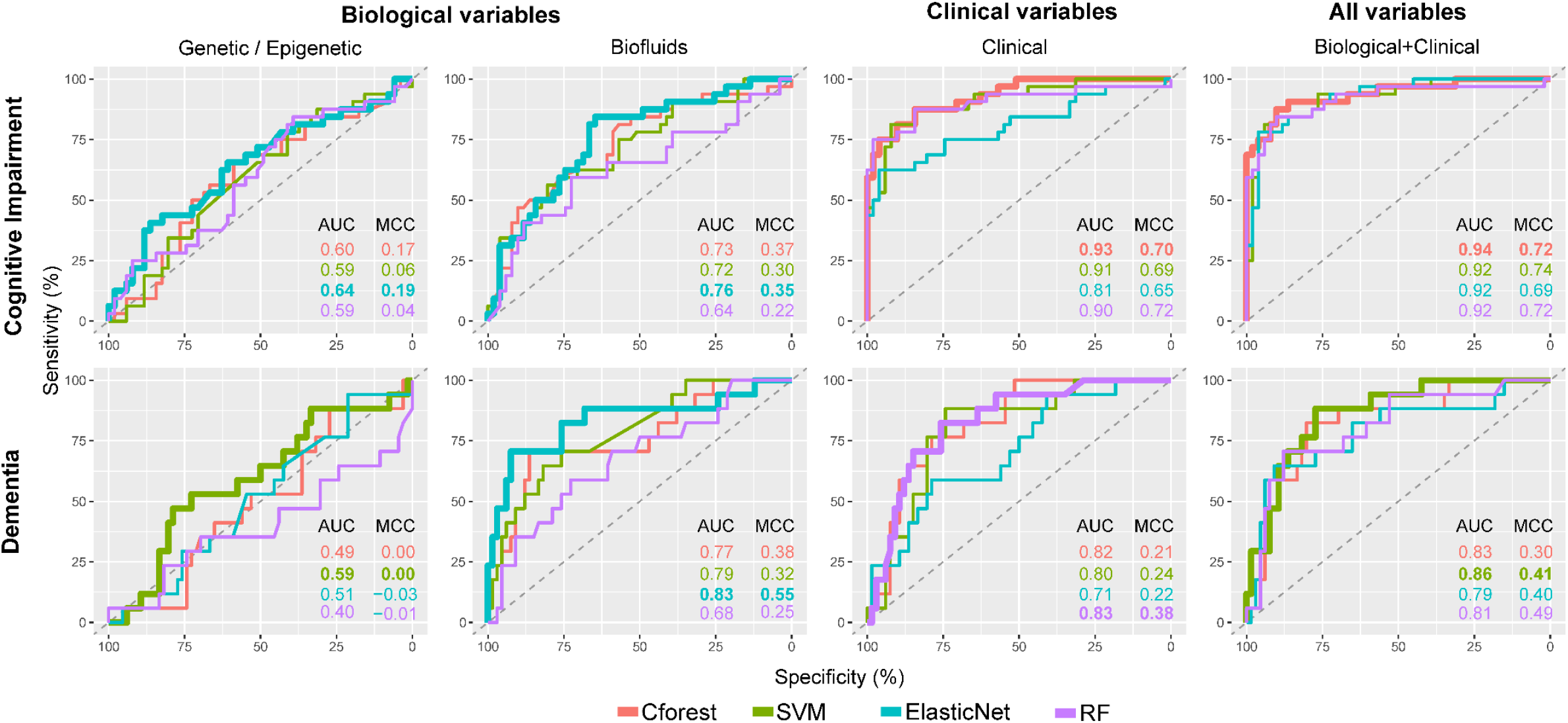
Receiver operating characteristic plots for predicting cognitive impairment and dementia using selected clinical, genetic/epigenetic and biofluid variables. ROC curves displayed in grid with rows as cognitive outcome and columns as variable subset. Colored by ML algorithm with the highest AUC for each outcome and variable set displayed as a thicker line. AUC and MCC metrics displayed as text for each plot. ROC: Receiver Operating Characteristic AUC: Area Under the Curve, MCC: Matthews Correlation Coefficient, ML: Machine Learning, SVM: Support Vector Machines, Cforest: Conditional Inference Random Forest, RF: Random Forest

Comparing both outcomes, prediction of cognitive impairment outcome showed better predictive accuracy than dementia conversion, reflected by higher area under the receiver operating characteristic curve (AUC) and Matthews Correlation Coefficient (MCC) metrics for all variable subsets. The one exception to this was biofluid measures, which when evaluating solely on AUC, appeared to show better prediction of dementia conversion than cognitive impairment. However, reviewing the prediction of dementia using biofluid measures shows poor overall prediction of true dementia converters when investigating MCC (Cforest = 0.38, SVM = 0.32, ElasticNet = 0.55, RF = 0.25) and sensitivity metrics (Table 2).

Overall, across both outcomes and variable sets, the best prediction was achieved for the cognitive impairment outcome using a combination of biological and clinical variables, reflected by high value balance for AUC and MCC (Table 2). This represented a marginal improvement over prediction of the cognitive impairment outcome using the clinical variable subset alone. Combining biological and clinical variable types improved sensitivity over the clinical models, represented by a higher number of true cognitive impairment predictions (Table 2).

The genetic/epigenetic variables alone showed minimal predictive accuracy irrespective of cognitive outcome and ML model tested, with near-random prediction, with AUC measures between 0.40 – 0.65 and MCC below 0.19 (Figure 2, Table 2).

### Predictive variables for cognitive impairment outcome

Given the best overall prediction was achieved using a combination of biological and clinical variables for the cognitive impairment outcome, we investigated individual variable contribution using Shapley values. Variables included by at least three ML algorithms are shown in Figure 3. Cognitive tests were heavily represented in overlapping models, with Hopkins Verbal Learning Test-Revised (HVLT-R) Immediate/Total Recall and Delayed Recall scores, Symbol Digit Modalities (SDM) and Semantic Fluency Test (SFT) being included in all four ML methods and Benton Judgment of Line Orientation (BJLO), HVLT-R Discrimination Score, Montreal Cognitive Assessment (MoCA), and SFT – Vegetable subscore being included in at least three (Figure 3A).

**Figure 3.**
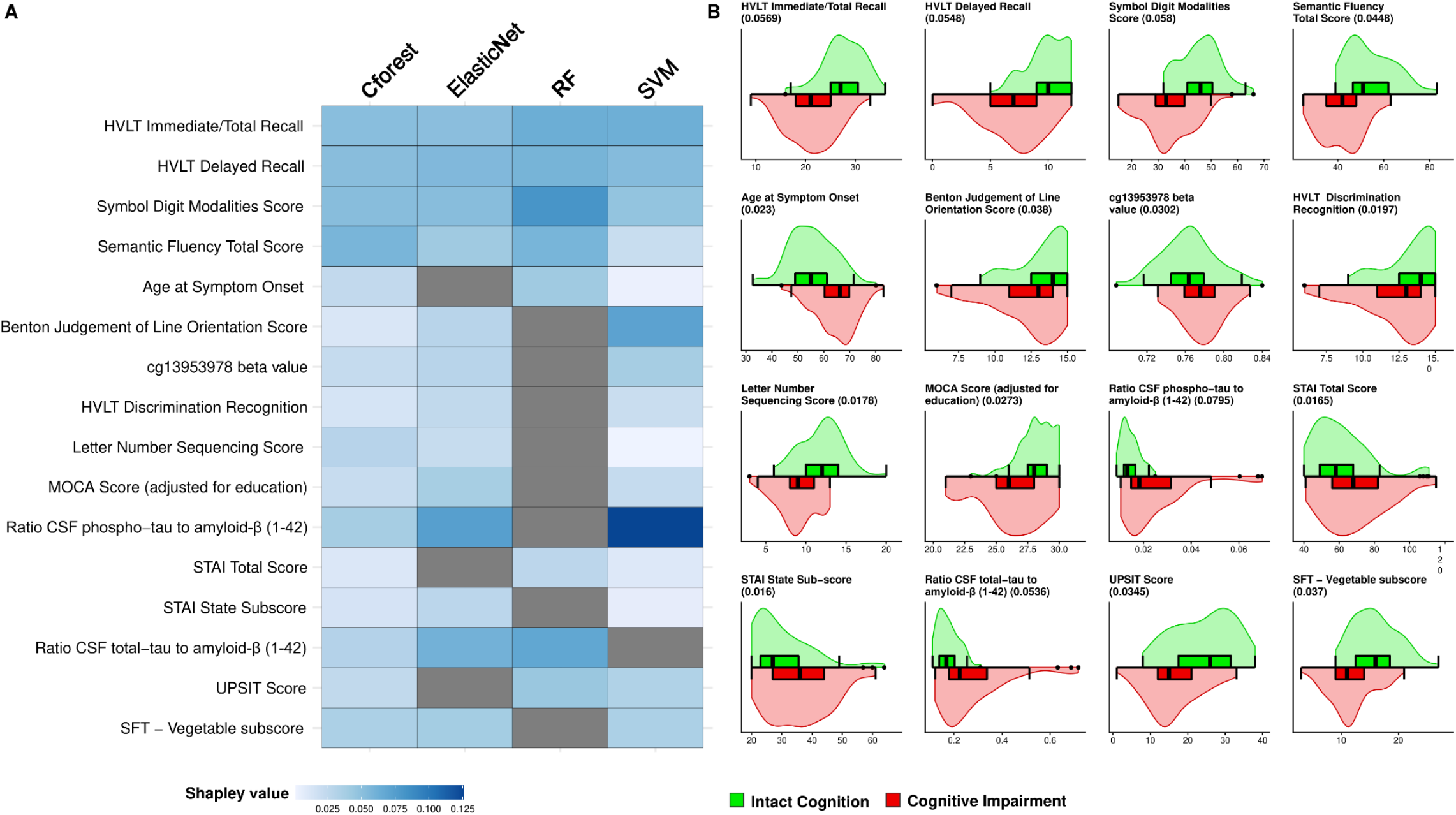
Variable importance in predicting cognitive impairment outcome. Variables included across three or more ML models for prediction of the cognitive impairment outcome using combined clinical and biological variables. **(A)** A heatmap of global Shapley importance. Darker blue reflects higher Shapley value and more important variables in the model. Variables not included in a particular model are shown in grey. **(B)** Dual violin and box plots of raw values of each variable between groups. Average global Shapley value importance for each variable is shown in brackets next to each variable name. Boxes represent median, Q1 and Q3 of the interquartile range (IQR) and whiskers display 1.5*IQR below and above Q1 and Q3 respectively. HVLT: Hopkins Verbal Learning Test, MOCA: Montreal Cognitive Assessment, CSF: Cerebrospinal Fluid, STAI: State-Trait Anxiety Inventory, UPSIT: University of Pennsylvania Smell Identification Test, SFT: Semantic Fluency Test, ML: Machine Learning.

Non-cognitive clinical measures included in multiple methods were age of symptom onset, State Trait Anxiety Inventory (STAI) scores (total and state sub-score) and the University of Pennsylvania Smell Identification test (UPSIT) for olfactory impairment. In these combined models, three biological variables showed consistently high contribution across multiple models including CSF Ratios of phospho-tau to amyloid-β (1-42) and total-tau to amyloid-β (1-42), respectively, as well as blood DNA methylation at cg13953978 (Figure 3A). Differences in overlapping variables are shown in Figure 3B, highlighting the direction of effect for each variable between cognitively intact and impaired groups.

Genetic variables were conspicuous in their absence from overlapping contributing variables, but were present in certain models, for example, *GBA* non-synonymous mutations were included for both Cforest and ElasticNet. Summarized Shapley value contribution across all tested algorithms are shown in Figures S1-4. As a graphical representation of prediction in our best performing model (Cforest), Figure S5 displays a surrogated decision tree, built by aggregating the best performing decision trees within the forest, containing a mix of biological and clinical variables. It is worth noting that this representation does not contain all variables included in the entire decision forest.

### The effect of cognitive tests in predictive accuracy

As we observed a large proportion of the top predictive variables were cognitive tests (9 out of 16, Figure 3A), we tested the sensitivity of predictions made without the use of cognitive variables. As Cforest models performed best on the clinical subset, we chose to explore the sensitivity of predictions with and without cognitive variables using this algorithm. Clinical variables were subset to cognitive only and non-cognitive variables as annotated in Table S1. As shown in Figure 4, we found that cognitive variables only (AUC = 0.90, MCC = 0.54) performed better than non-cognitive variables (AUC = 0.86, MCC = 0.46). The combination of the two variable subsets into an overall clinical model showed a marginal increase in AUC (0.90 to 0.93) but a larger increase in sensitivity reflected by increased MCC from 0.54 to 0.70.

**Figure 4.**
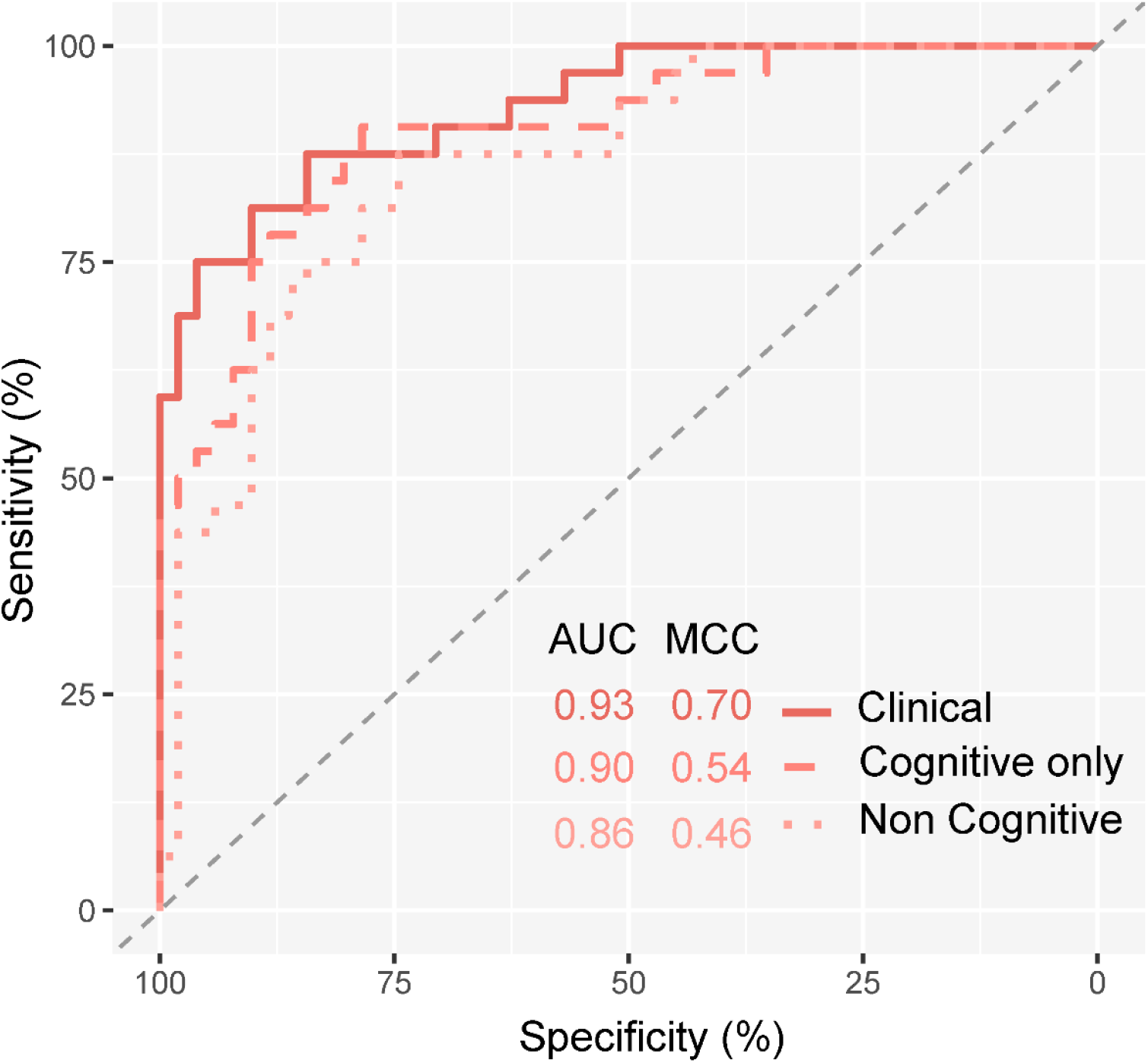
Sensitivity analysis of cognitive variables. ROC showing prediction of the cognitive impairment outcome using Cforest applied on clinical subsets. Non-cognitive variables: light grey dotted line, cognitive variables: dashed grey line, all clinical variables: solid black line. Summary of AUC and MCC metrics for each subset shown in plot text. AUC: Area Under the Curve, MCC: Matthews Correlation Coefficient, Cforest: Conditional Inference Random Forest, ROC: Receiver Operating Characteristic

## Discussion

In the present study, we tested the prediction of two cognitive outcome measures in newly diagnosed PD subjects within eight years, using multiple variable subsets and ML algorithms. The generated models were assessed for metrics of prediction accuracy and the importance of contributing variables. We found that combining both biological and clinical variables produced best performing models, with a marginal improvement in predictive performance compared to models using clinical variables alone. We interpret this as evidence of synergistic contribution of multivariate data types, producing the most accurate predictions. Of variable subsets, the most accurate and balanced prediction was achieved when testing for cognitive impairment (MCI or dementia) using clinical data, giving the highest AUC, MCC metrics and balance of sensitivity and specificity. When evaluated individually, non-clinical measures (biofluids and genetic/epigenetic) showed poor predictive performance, regardless of outcome tested and ML algorithm used.

Comparing outcomes, prediction of cognitive impairment consistently outperformed dementia conversion, which we interpret as being driven by poor differentiation of MCI individuals when predicting dementia conversion. MCI is a well-established risk factor for future dementia development^4^, and previous studies show higher dropout within PPMI is associated with worse cognitive performance^19^. Given this, the overall progression profile of MCI and dementia, as subsets within this study, might not differ substantially, with MCI patients potentially converting to dementia in unobserved events. This further supports the use of a combined cognitive impairment group, with best prediction being observed for this outcome.

Unsurprisingly a high number of contributing variables included cognitive assessments, indicating that there was already a level of cognitive changes present at baseline. This highlights a potential limitation in the inclusion of these variables, as these cognitive assessments are highly associated with the outcome of interest we aimed to predict. However, these measures reflect an assessment time 1-7 years before a clinically diagnosed conversion to either MCI or Dementia. Sensitivity analysis of the effect of cognitive variables in prediction confirmed that cognitive variables had a large contributory effect to predictions although increased sensitivity was observed with the inclusion of non-cognitive clinical variables. Top contributing non-cognitive variables included age at onset of PD, anxiety, and olfactory impairment. Older age of PD onset, which we observe within the cognitive decline group, is a well-established and validated risk factor for PD-cognitive decline^4^. Olfactory impairment has been increasingly associated with cognitive impairment in PD^20-23^. Although anxiety is less associated as a predictive variable for cognition within PD^24^, it has been associated as a predictor of worse cognitive prognosis in general population studies^25^.

Within combined models utilizing both biological and clinical variables, ratios of CSF protein measures of total-tau, phospho-tau and amyloid-β (1-42), had a high contributory effect across multiple ML algorithms. Additionally, one measure of blood DNA methylation, cg13953978, was included in multiple combined models. This locus has been previously associated with multiple neurodegenerative diseases and, of note, we observe the same direction of effect between cognitively impaired and preserved individuals in this study and previously reported findings^26^.

Several studies have aimed at creating an accurate model to predict cognitive outcome in PD using the PPMI cohort^9,16,27^. Compared to previous studies, in the current study, we have included a larger range of biological variables including polygenic scores for multiple related traits and epigenetic measures. We used MDS criteria for defining cognitive performance at each follow-up as a substitute for the commonly used MoCA. Additionally, we included a long follow-up period and excluded reverters from the modelling.

To improve the accuracy and generalizability of our models compared to other models reported previously, we employed a multi-objective model optimization procedure using three criteria (AUC, MCC, and number of variables). Although AUC is commonly used for model interpretation, it is insensitive to class imbalance. Therefore, to prevent inaccurate prediction assessment, we included MCC, as this metric can evaluate accuracy while considering class balance. This, along with recursive feature elimination (RFE)^28,29^ and k-fold cross validation, further avoided the risk of overfitting and addressed the high number of variables included in this dataset. We applied multiple ML algorithms, to cover a range of different learning strategies, standardly applying RFE and multi-objective optimization for each.

A potential limitation of this study is the curatorial nature in which cognitive groups were subset and the relatively small sample size available. We justify the methods for cognitive group subsetting as we aimed to represent individuals with clinically relevant diagnoses confirmed by multiple observations over time. However, due to data missingness and attrition within PPMI, there are a number of de-novo cases enrolled at baseline which were not tested within our models.

Our findings of DNA methylation at cg13953978 as a predictive variable requires further replication to ensure it is not the result of an unknown cryptic stratification in this cohort. Previous association of this loci with neurodegenerative disease across multiple cohorts do however support it as a potential biomarker. Expanding the number of genetic and epigenetic variables included in future studies to a genome-wide level in cohorts designed around cognitive decline prediction is also essential to truly uncover potential predictive efficacy. However, due to the challenge of including the high number of variables implicit in multi-omics data^19,30-32^, we found this to be outside of the scope of the current study.

In summary, after evaluating multiple predictive variable types and outcomes, we established a model that accurately predicted cognitive impairment and preserved normal cognition over a follow up eight-year time span. This prediction was largely driven by clinical measures of both known risk factors and more novel measures, but also variably included biological variables. This work supports evidence of anxiety and olfactory impairment as potential predictors of cognition in PD and highlights epigenetic measures of DNA methylation as biological predictive variables requiring further investigation.

## Methods

### Participants and cognitive assessment

All data used in this study was obtained from the PPMI^18^ database (https://ida.loni.usc.edu/). Participants were selected from the de novo PD cohort, defined by a diagnosis of the disease within two years and un-medicated for motor symptoms at baseline (n = 423). Subjects underwent yearly cognitive diagnosis in accordance with Movement Disorders Society (MDS) recommended criteria for dementia and MCI as previously reported^19–21^. In brief, a confirmed MCI diagnosis was based on an impaired performance on at least two test scores > 1-2 standard deviations below a standardized mean^33^. Dementia diagnosis alongside clinical annotation required impaired performance in at least two cognitive domains coinciding with significant functional impairment resulting from cognitive state^34^.

Records of cognitive diagnoses from baseline to year eight were sourced from PPMI following their routine application of the above criteria to create three groups of PD patients with distinct cognitive outcomes as follows (Figure 1 and S6):

#### PD-Dementia

Cases showing any diagnosis of dementia over an eight-year time span were annotated as the dementia conversion cases, excluding one individual that reverted to normal cognition after an annotation of dementia (n = 43).

#### PD-MCI

Cases with any record of MCI without any annotation of future dementia diagnosis (n = 39) were annotated as PD-MCI conversion cases. This group excludes a set of 14 cases that reverted to normal cognition following MCI annotation.

#### Cognitively Intact (CI)

To avoid any effect of attrition and cognitive decline in unobserved events, cases defined as cognitively intact required a minimum of five records of normal or subjective cognitive decline (SCD) during recorded visits up to year eight (n = 127). This excluded 175 cases showing missing values or indeterminate diagnoses.

Subsequently, we used these groups to define two separate binary outcomes for machine learning-based prediction as follows:

#### Cognitive Impairment Outcome

Defining conversion to cognitive impairment within an eight-year time span. This compared the CI group (n = 127) to an impaired group, created by combining the PD-Dementia and PD-MCI groups (n = 82).

#### Dementia Conversion Outcome

Defining conversion to dementia within an eight-year time span. This compared the PD-Dementia group (n = 43) to a non-dementia conversion group created by combining PD-MCI and CI groups (n = 166).

### Baseline data

Baseline data for all 423 PD were sourced from PPMI and processed into four sets of variables (Table S1):

#### Clinical variables

These included demographic variables (gender, age of onset, years in education, duration of disease, family history of PD), motor symptoms (MDS-UPDRS Part 1 and 2 total scores, rigidity score, tremor dominant / postural gait instability disorder classification, Hoehn and Yahr [H&Y] scale, Modified Schwab & England Activity Daily Life [ADL] Score), psychiatric symptoms (MDS-UPDRS Part 1 subscores, Geriatric Depression Scale [GDS], Questionnaire for Impulsive-Compulsive Disorders, State Trait Anxiety Test), autonomic symptoms (SCOPA-autonomic subscores), sleep disorder (Epworth Sleepiness Scale Score [ESS], Categorical REM Sleep Behavior Disorder Questionnaire subscore, MDS-UPDRS Part 1 subscores) and olfactory symptoms measured by University of Pennsylvania Smell Identification Test (UPSIT). Assessments of cognition (Semantic Fluency Test [SFT], Symbol Digit Modalities [SDM], MDS-UPDRS Part 1 subscores, Montreal Cognitive Assessment [MoCA], Hopkins Verbal Learning Test-Revised [HVLT-R] subscores, Benton Judgment of Line Orientation [BJLO]) were also included.

#### Biofluid variables

CSF measures for amyloid-β (1-42), phospho-tau181, total-tau and α-synuclein were included, after removing cases showing high levels of CSF hemoglobin (> 200 ng/mL) as previously described^35,36^. Ratios of each measure were also included as independent predictive variables. Total serum uric acid was also included as previously described^37^.

#### Genetic and Epigenetic variables

Genetic variables included individual *APOE* genotype, MAPT haplotype and the SNPs rs12411216^38^, rs356181^39^ and rs3910105^40^. *GBA* mutation status was included as a binary factor for the presence of any non-synonymous coding mutations present within the *GBA* region. Polygenic risk scores (PRS) were calculated for Parkinson’s disease (*GBA* region excluded), Alzheimer’s disease (AD) (*APOE* region excluded), educational attainment (EA), schizophrenia (SCZ), major depressive disorder (MDD) and coronary artery disease (CAD) (Supplementary Methods).

After stringent quality control and normalization of the whole-genome DNA methylation data measured in baseline blood (Supplementary Methods), 21 loci were selected based on previously reported differentially methylated positions associated with cognitive decline in PD^41^ or across neurodegenerative disease^26^. Epigenetic age acceleration measures from the GrimAge clock^42^, BloodAndSkin clock^43^ and the modified Hannum clock which included measures of both intrinsic epigenetic age acceleration (IEAA) and extrinsic epigenetic age acceleration (EEAA, incorporating intrinsic measures as well as blood cell proportions)^44^ were included as additional epigenetic variables.

#### Combined biological and clinical variables

This variable set collated all previously listed variables across the clinical, biofluid and epigenetic/genetic subsets into one combined total set.

Summary lists of measures used for predictive modelling are shown in Table S1 and descriptive statistics in Table 1. All measures highlighted in this summary table were carried forward for multivariate modelling.

### Data Processing

#### Imputation

Each baseline variable was evaluated for the proportion of missing observations and missing values imputed using available data for the selected variable. For ordinal and categorical variables, the mode value was chosen for imputation, for continuous variables the median value was selected.

#### Stratification

Due to an imbalance in the size of selected outcome groups, stratified sampling was used to account for potential training imbalance and testing bias^45^ using the ‘*stratified*’ function from the *splitshapestack* R package (version 1.4.8). Sampling considered the proportion of outcome groups, the proportion of MCI and dementia cases as well as gender and categorical age (1: <56 years, 2: 56-65 years, 3: >65 years). A 60/40 train/test split was chosen to increase samples in the test set to give an improved evaluation of the final resulting models.

#### Data transformation

The baseline data contains three types of variables: categorical, ordinal, and continuous. To ensure each variable had a similar influence during the ML process, Z-score normalization was performed using the base R function ‘*scale’* on the continuous variables based on averages of the training set^46,47^. The parameters ‘center’ and ‘scale’ were stored per variable and used to re-scale the training and testing data accordingly.

### Machine Learning

#### Training and selected algorithms

The R package *caret* (version 6.0.90) was used to establish the machine learning workflow and tune the hyperparameters^48^. We used four different classifiers from three machine learning families. The selected algorithms include functions for RF (‘*rf’*) and conditional inference forest (‘*cforest’*) from the RF family, SVM with linear Kernels (‘*svmLinear’*) from the support vector machine family and ElasticNet (‘*glmnet’*) from the generalized linear model family of classifiers. RF and Cforest are information-based learning algorithms, and their behavior is determined by concepts from information theory^49^. RF algorithms are based on a majority vote of a collection of different decision trees. Cforest differs from RF as it does not select variables based on maximization of an information measure but based on a permutation test for significance^50^. SVM and ElasticNet are error-based learning algorithms, and their behavior is explained by minimizing total error during training^49^. SVM algorithms are based on generating the best possible separation between classes of interest in a hyperdimensional pane. ElasticNet is a generalized linear model with L1 and L2 regularization, able to shrink or drop coefficients to achieve a better model fit.

#### Tuning

To avoid overfitting during training, 10 repeated 10-fold cross-validation was used. During the training process, hyper-tuning was enabled with a maximum of 100 tunes to promote model accuracy. To prevent optimistically inflated results due to imbalanced datasets, we used MCC alongside AUC to evaluate model accuracy^47,51^.

#### Variable selection and model generalization

Recursive feature elimination (RFE) was applied as the variable selection algorithm. In brief, RFE iterates through generations of models using a decreasing training set, eliminating the worst contributing variable of each iteration^52^. The first model was trained using all available variables, with the resulting evaluation metrics being extracted and stored. Variable importance was recursively calculated for the generated model using the ‘*varImp’* function in *caret*. The least contributing variable was flagged to be removed in the next iteration. The updated training data was used to train a new model, and the process was repeated until one variable remained. This resulted in numerous models with decreasing number of variables.

#### Optimal model selection

To reduce generalization error, a multi-objective optimization procedure was applied by utilizing MCC, AUC and the number of variables from each model in each iteration^53^. MCC and AUC were chosen as MCC is calculated on binary classes while AUC is calculated by class probability, allowing model selection to benefit from the properties of MCC and the resolution of AUC. This ensures model generalization with higher accuracy. Moving averages of these metrics (window = 5) were calculated and the rank was determined (Figures S7 and S8). Calculating the mean rank of the moving averages allows a comparable scale to the variable number per each *i-th* model. From this we calculated an optimal model score by adding together the number of variables to the average rank, as shown in the formula below. This results in an optimization curve highlighting the best performing model with the lowest number of variables. The model with the lowest score was selected as the optimal model, as this model indicates the highest accuracy, balanced prediction, and least number of variables.

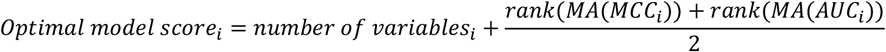

#### Testing

The optimal model was used for class prediction on the test dataset, yielding several evaluation metrics (AUC, MCC, Accuracy, Sensitivity, Specificity) as well as other evaluation elements (such as confusion matrices, Receiver Operator Characteristics (ROC)-AUC curves, and individual variable difference plots).

#### Variable importance calculation

Shapley values were calculated for the interpretation of individual variables included in best performing models. Using the package *iml* (version 0.10.1), a predictor object was generated, containing the model of interest and the test dataset. This predictor object was used in the calculation of the Shapley values per sample, with 10,000 Monte-Carlo-Simulations. The resulting absolute Shapley values were averaged over all samples, yielding global Shapley contribution per variable^54^.

## Data Availability

Data used in the preparation of this article were obtained from the Parkinson's Progression Markers Initiative (PPMI) database (www.ppmi-info.org/access-dataspecimens/download-data). For up-to-date information on the study, visit ppmi-info.org.
All codes are available at https://github.com/Rrtk2/PPMI-ML-Cognition-PD

## Code availability

All codes are available at https://github.com/Rrtk2/PPMI-ML-Cognition-PD

## Data availability

Data used in the preparation of this article were obtained from the Parkinson’s Progression Markers Initiative (PPMI) database (www.ppmi-info.org/access-dataspecimens/download-data). For up-to-date information on the study, visit ppmi-info.org.

## Contributions

E.P. conceived and directed the project. J.H. and R.A.R. undertook data analysis, and support with data review. J.H., R.A.R. and E.P. wrote the first draft of the manuscript. A.D. and B.C. were involved in the selection of the clinical predictors and outcome. R.C., S.K., A.T. provided advice on data analysis. G.S. contributed to generating polygenic scores. J.H., R.A.R., E.P., K.L., A.F.G.L., L.E., B.P.F.R., B.C., A.D. contributed to the interpretation of the results. All authors provided critical feedback on the manuscript and approved the final submission. PPMI – a public-private partnership – is funded by the Michael J. Fox Foundation for Parkinson’s Research and funding partners, including [list the full names of all of the PPMI funding partners found at www.ppmi-info.org/about-ppmi/who-we-are/study-sponsors].

## Notes

### Competing Interest Statement

The authors have declared no competing interest.

### Funding Statement

ZonMw (Netherlands Organisation for Health Research and Development) - 733050516 [Pishva]

### Author Declarations

This study involves only openly available data from the PPMI study https://www.ppmi-info.org/

