## Supplementary Materials for "Machine learning-based prediction of cognitive outcomes in de novo Parkinson’s disease"

### Supplementary methods

##### *Genotyping and polygenic scores calculation*

Whole blood DNA genotyping was previously performed on the NeuroX SNP array by PPMI investigators using published methods[1]. Raw data from 423 individuals covering 267,607 variants was quality control (QC) assessed following published recommendations[2]. In brief, data was excluded based on the following criteria 1) variants and individuals with missingness >0.1, 2) individuals with discordant reported sex and inferred sex (X chromosome homozygosity F-value >0.8 for males, <0.2 for females), 3) variants with minor allele frequency <0.01 or >0.05, 4) variants deviating from Hardy Weinberg Equilibrium < 1e-3, 5) individuals with heterozygosity rate +/- 3 standard deviations, 6) individuals with evidence of cryptic relatedness (pi hat >0.2). Following initial QC, autosomal data was extracted, plink files were converted to vcf format and uploaded to the Michigan Imputation Server. Imputation was conducted using Eagle2 to phase haplotypes and Minimac4 using the 1000 Genomes reference panel (phase 3, version 5). An R2 filter score for imputation quality was set at 0.3. Following imputation, data was downloaded, converted to plink format and quality assessed following the previous criteria. Finally, genetic principal components were generated along with reference data from the 1000 Genomes Project and non-European cases removed based on qualitative assessment of clustering of the first two principal components. 582 cases passed QC (total variants post-imputation n = 2,287,446).

Polygenic risk scores (PRS) were calculated using summary statistics from recent genome wide association studies (GWAS) for Alzheimer's disease (AD) [3], Parkinson's disease (PD)[4], education attainment (EA)[5], schizophrenia[6] and major depressive disorders[7]. For AD, the effect of the *APOE* region was excluded by removing the region chr19:45,116,911 – chr19:46,318,605. For PD, the effect of the *GBA* region was removed by removing. PRSice-2 software[8] was used for polygenic risk score calculation, which automates clumping and p-value thresholding to generate a "best-fit PRS" for a target phenotype of interest. Briefly, clumping was performed to retain the most significant GWAS variants in a linkage disequilibrium (LD) block (250kb window, r2 threshold = 0.1). The PRS model is tested over an increasing set of p-value threshold (5e-08 to 1), with the optimal threshold set which generates a score explaining the maximum phenotypic variance in the target phenotype of interest. Phenotype was coded as a binary factor of 0 (Control) and 1 (PD) for this analysis, with the first eight genetic principal components used as covariates[9].

***DNA methylation data processing***

Whole blood genome-wide methylation in the PPMI cohort at baseline has been previously profiled on Illumina EPIC Array at baseline has been previously reported[10]. These included individual previously associated methylated loci as well as epigenetic age prediction features. Raw IDAT files were downloaded from the PPMI database (https://ida.loni.usc.edu/) in April 2020 and processed using the R package *wateRmelon*[11]. For epigenetic age prediction and age acceleration analysis non-normalized beta values were uploaded to the web-based tool https://dnamage.genetics.ucla.edu, selecting the "normalize data" and advanced analysis" options. For inclusion of specific epigenetic loci, data was quality controlled and normalised following established pipelines. Briefly, samples with low signal intensities or bisulphite conversion rate, mismatched reported and imputed sex or cryptic relatedness were excluded. P-filtering was applied using the '*pfilter'* function in the *watermelon* package, excluding samples with >1 % of probes with a detection P value >0.05 and probes with >1 % of samples with detection P value >0.05. Beta values for each probe were quantile normalised using the '*dasen' function*.

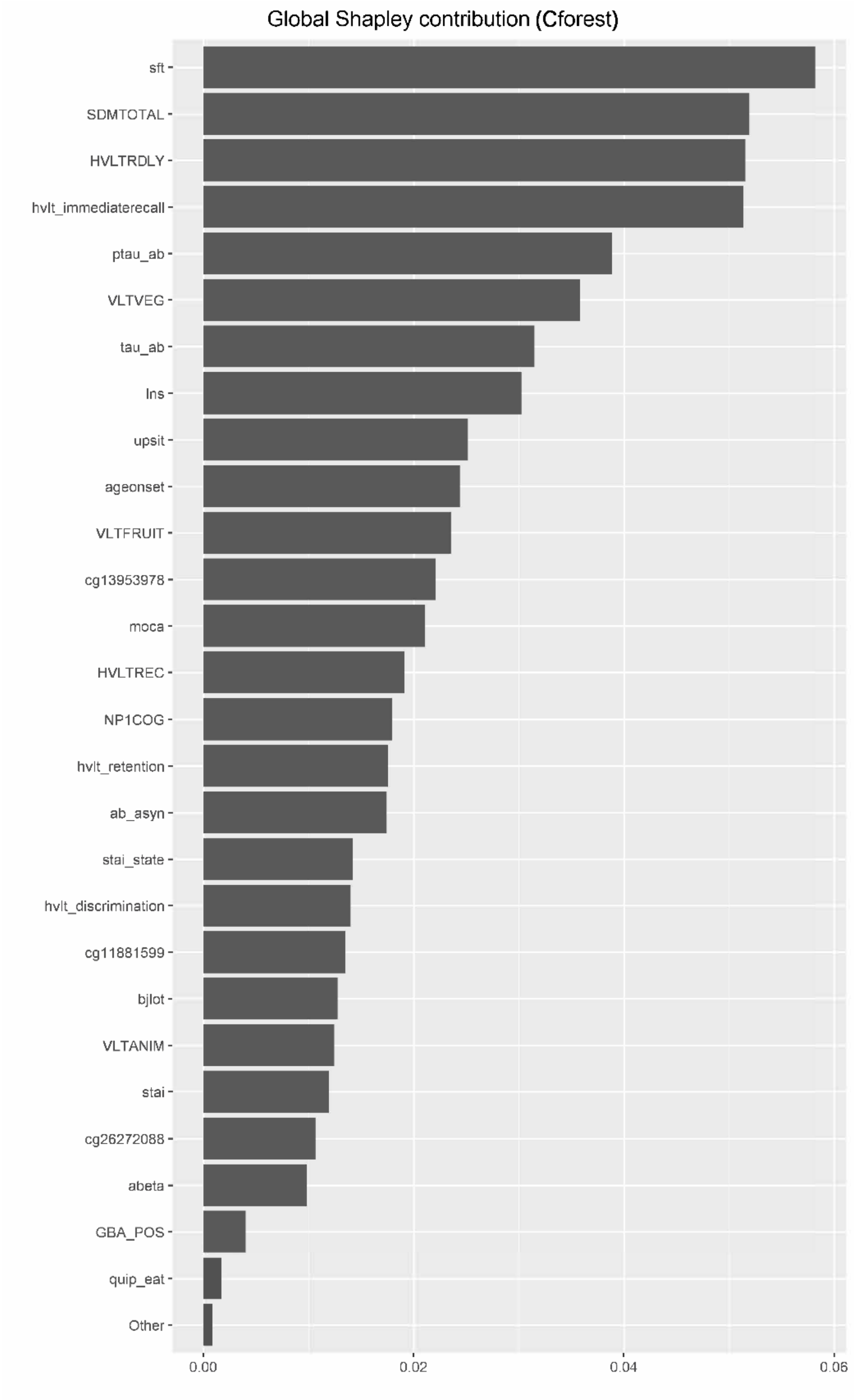


**Figure S1 Summary of variables included in Cforest ML prediction of the cognitive impairment using combined clinical and biological variable set.** Raw Shapley values are shown as bar values. Variable short names are listed in Table S1. The largest bars are at the top of the figure and represent higher Shapley value and more important variables in the model.

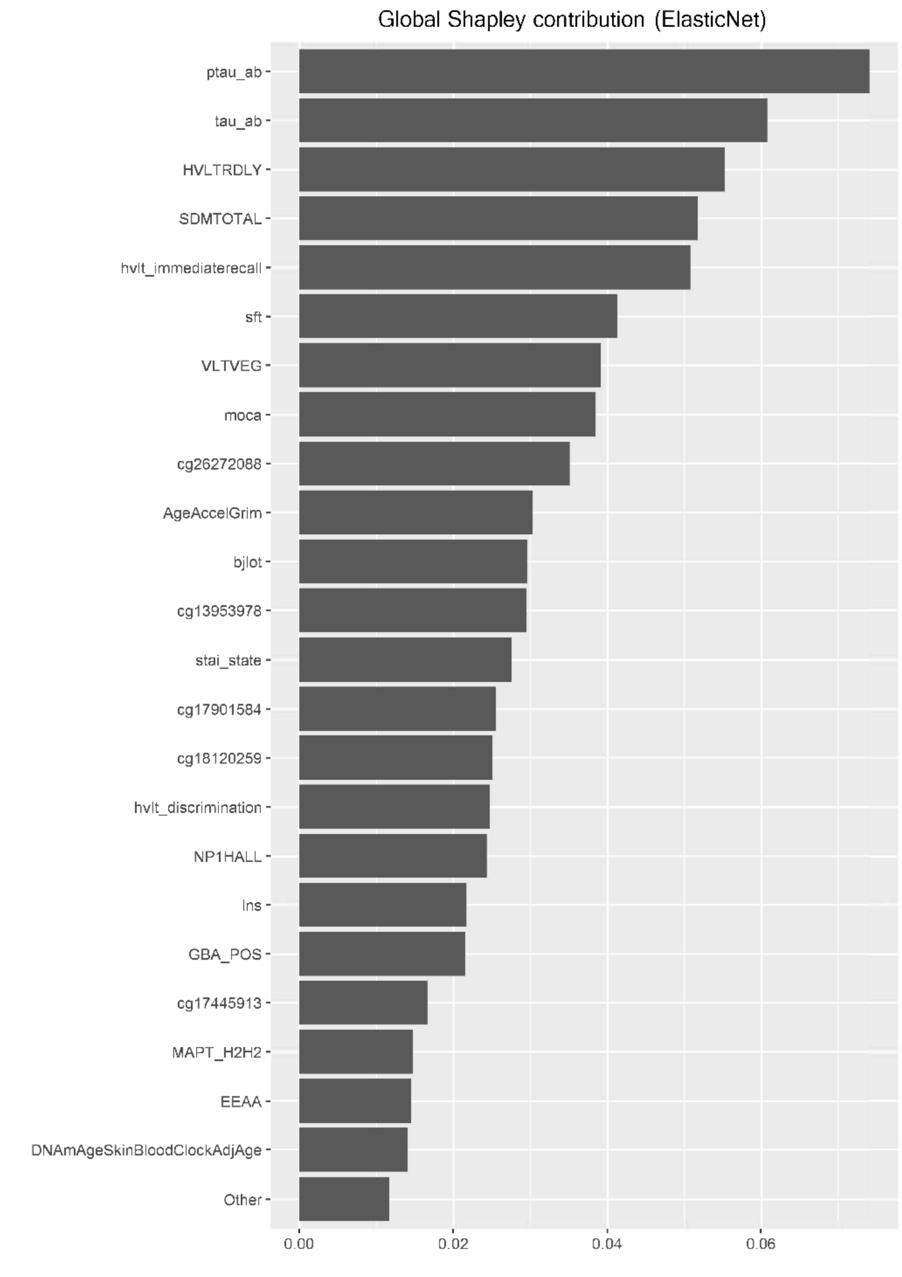


**Figure S2 Summary of variables included in ElasticNet ML prediction of the cognitive impairment using combined clinical and biological variable set.** Raw Shapley values are shown as bar values. Variable short names are listed in Table S1. The largest bars are at the top of the figure and represent higher Shapley value and more important variables in the model.

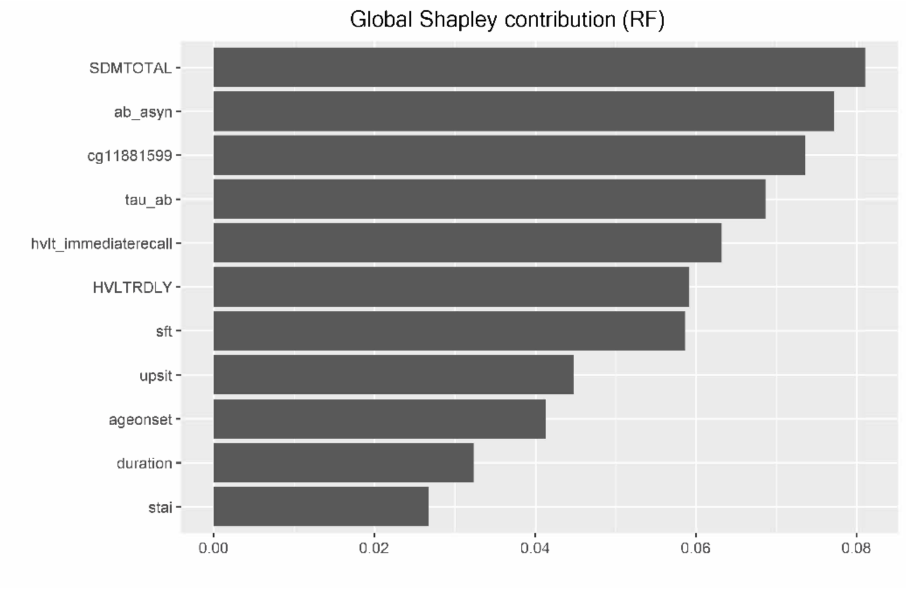


**Figure S3 Summary of variables included in RF ML prediction of the cognitive impairment using combined clinical and biological variable set.** Raw Shapley values are shown as bar values. Variable short names are listed in Table S1. The largest bars are at the top of the figure and represent higher Shapley value and more important variables in the model.

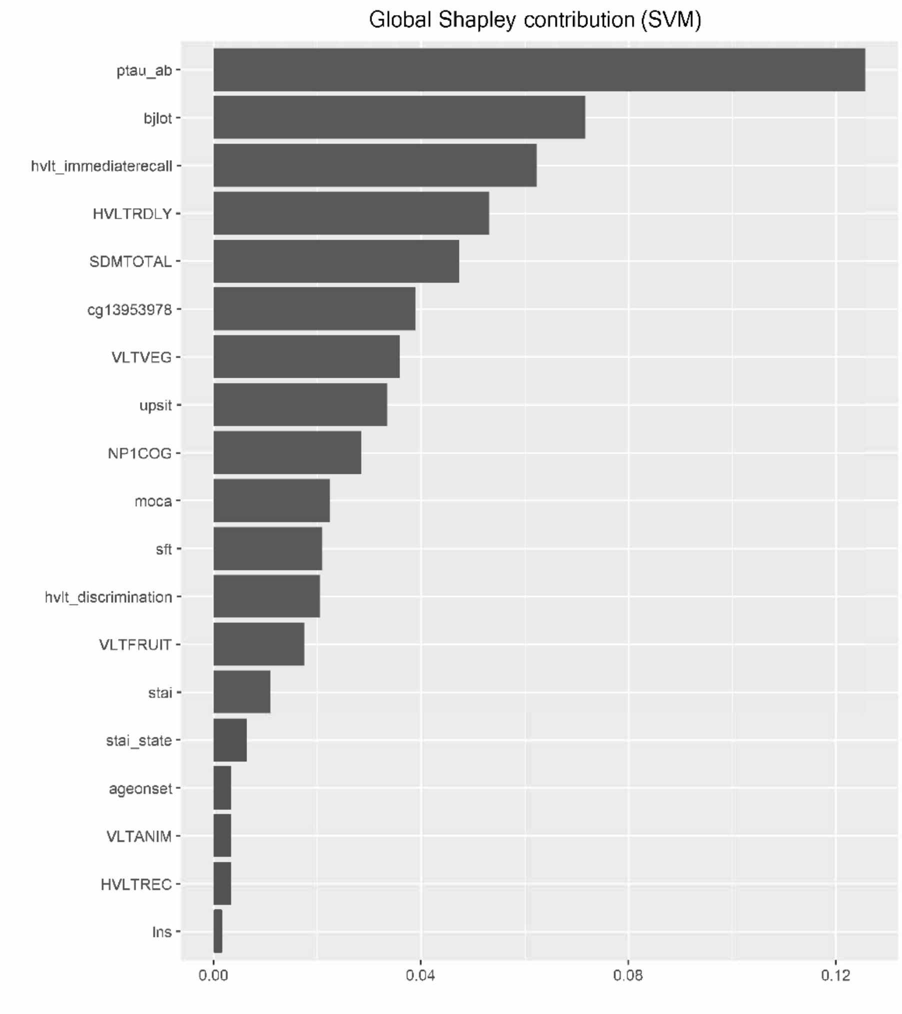


**Figure S4 Summary of variables included in SVM ML prediction of the cognitive impairment using combined clinical and biological variable set.** Raw Shapley values are shown as bar values. Variable short names are listed in Table S1. The largest bars are at the top of the figure and represent higher Shapley value and more important variables in the model.

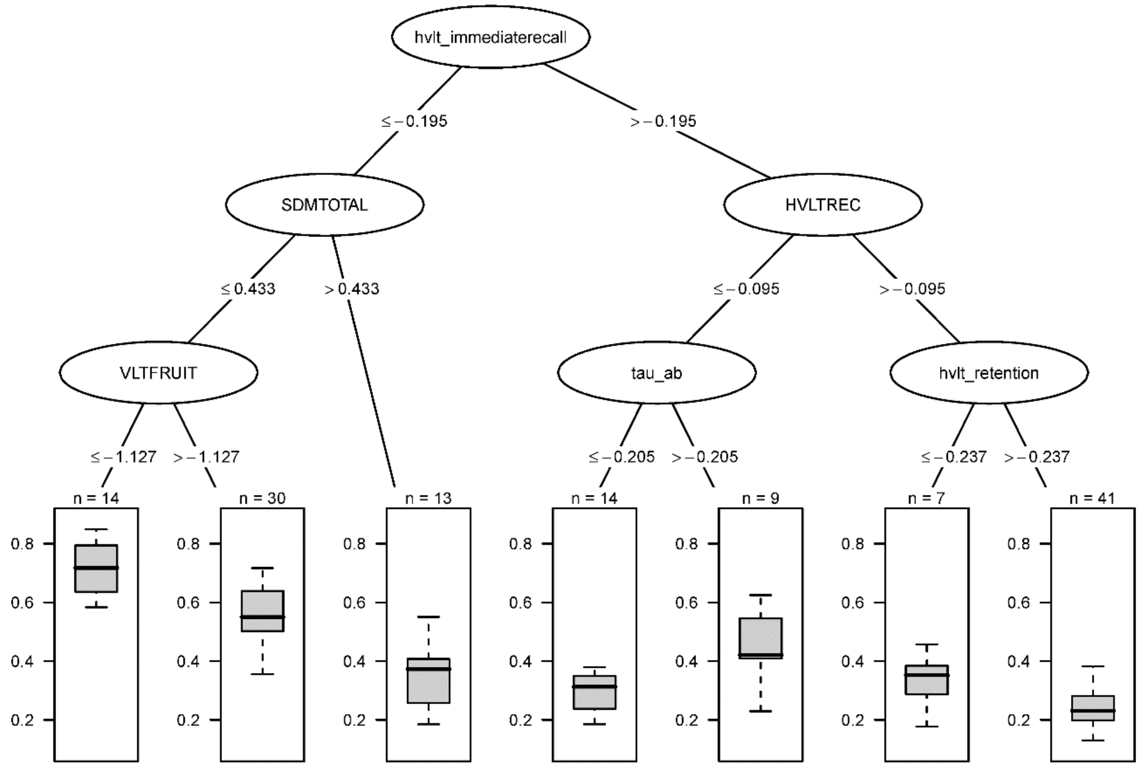


**Figure S5 Representative surrogate tree generated from full Cforest.** Shown is a representative surrogated tree approximating the full forest. Nodes indicate the variables (short name as outlined in Table S1), lines between nodes indicate the decision with corresponding criteria (Z-score normalized values, see methods), boxplots indicate probability of developing cognitive impairment. Variables are ordered on discrimination power, the first node to evaluate is hvlt_immeriaterecall, and based on the outcome the next node needs to be evaluated. Sequential decisions will result in a probability range of developing cognitive impairment shown in bottom boxplots. Number of classified training samples shown above each boxplot.

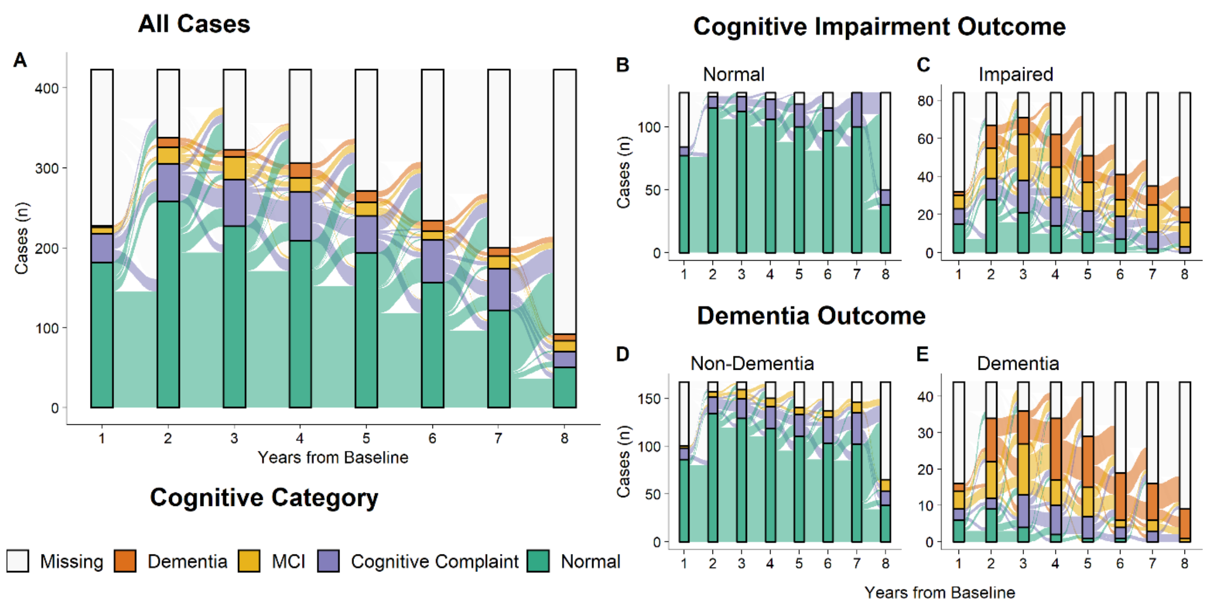


**Figure S6 Alluvial plots showing individual proportions of cognitive diagnosis for each yearly visit. The individual change in cognitive diagnosis between years represented by flow lines between nodes**. (A) All enrolled de-novo PD cases in PPMI (n = 423). Subset groups for each outcome measure shown on the right-hand side. (B, C) Subset groups for the Cognitive Impairment Outcome corresponding the cognitively intact group (n = 127) and the impaired group (n = 82). D, E) Subset groups for the Dementia Conversion outcome including the non-Dementia converting group (n = 166) and the Dementia converting group (n = 43).

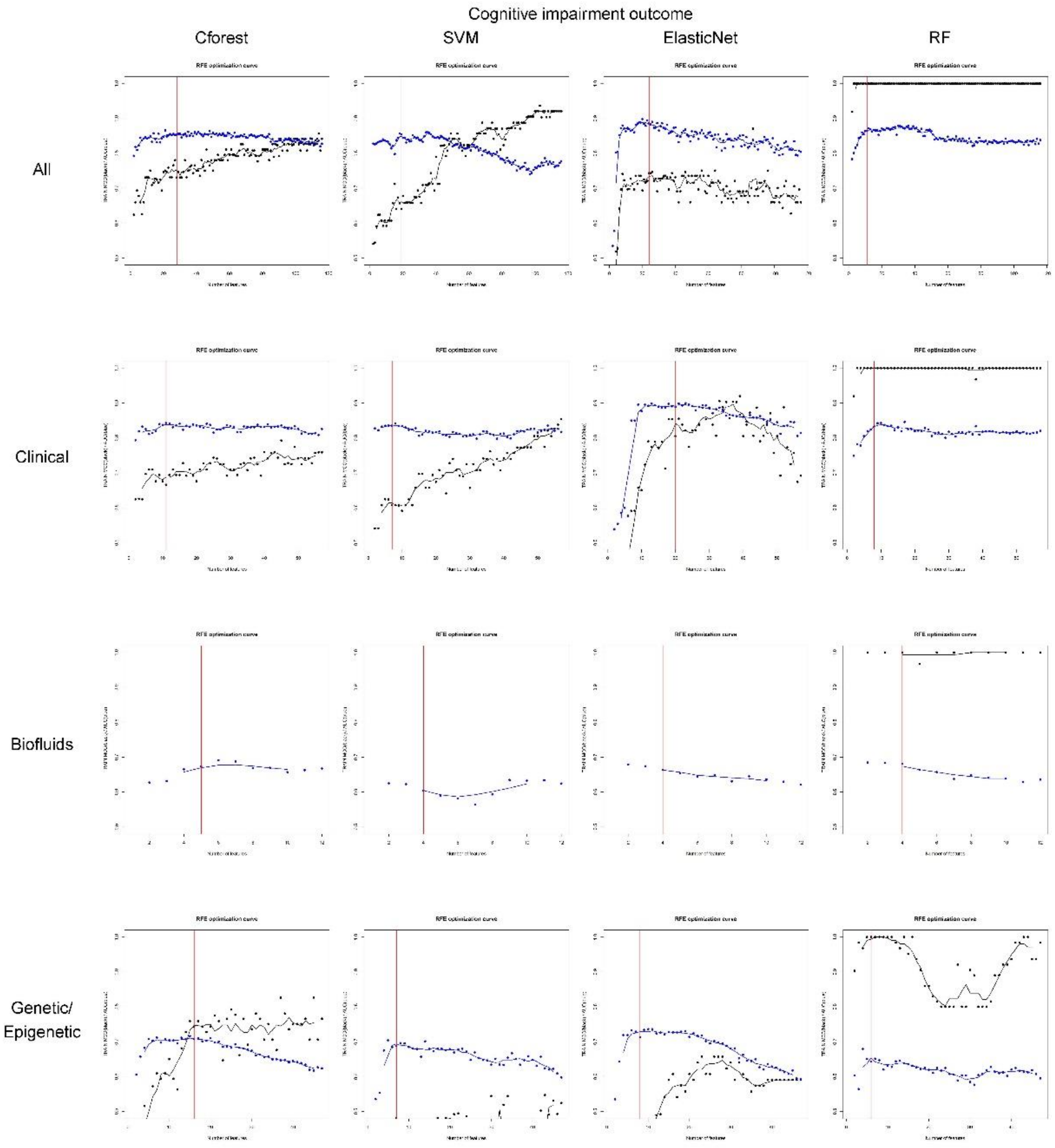


**Figure S7 Individual optimization curves for Recursive Feature Elimination for Cognitive Impairment Outcome.** The number of variables added per iteration is shown along the x axis and accuracy measurements on the y axis. AUC is shown in blue and MCC in black, with lines indicating the moving average. A red line on each plot indicates the optimum model according to multi objective optimization, as detailed in the methods. Plots are organized in rows for variable subsets and columns for ML algorithms.

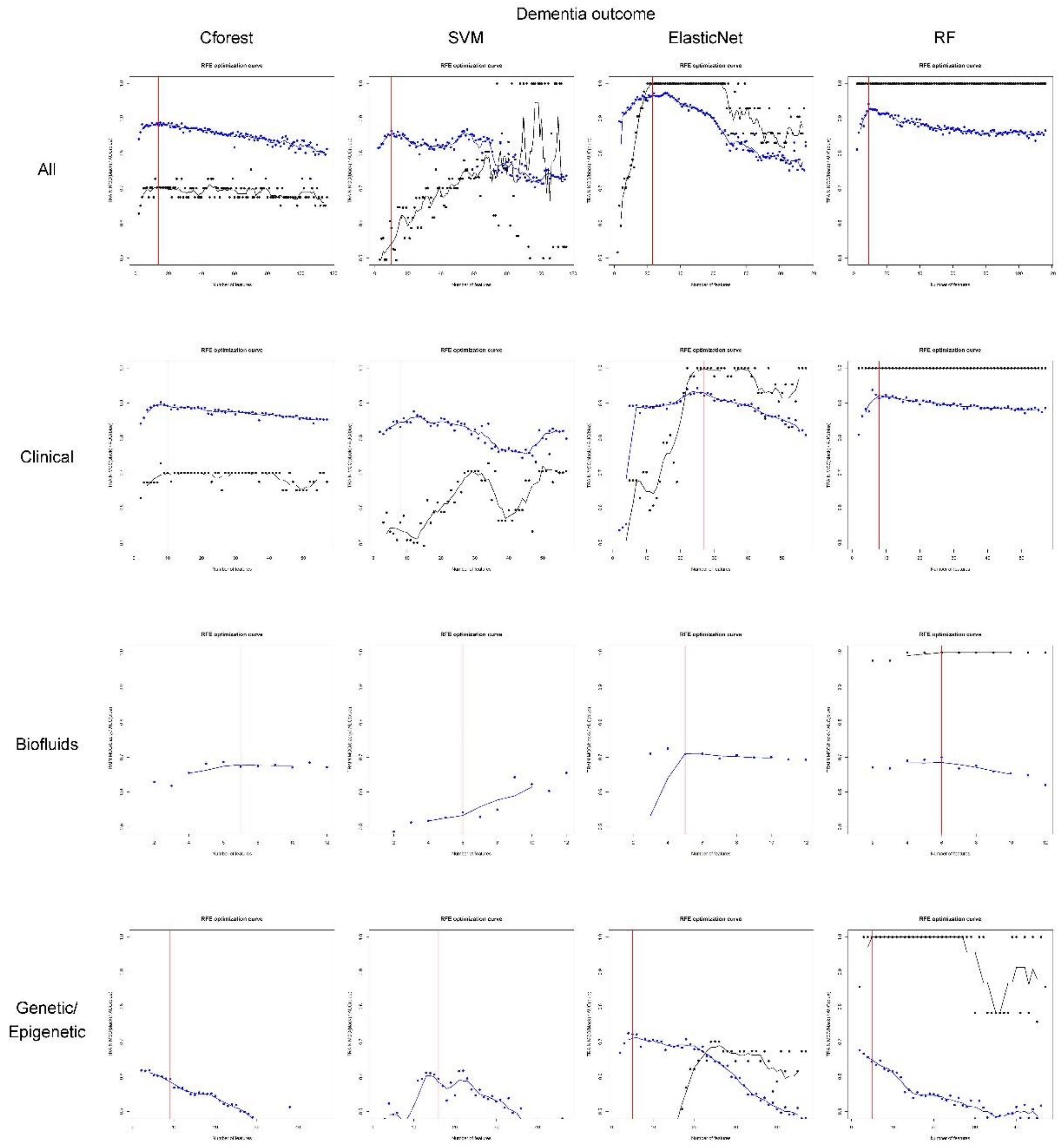


**Figure S8 Individual optimization curves for Recursive Feature Elimination for Dementia Conversion Outcome.** The number of variables added per iteration is shown along the x axis and accuracy measurements on the y axis. AUC is shown in blue and MCC in black, with lines indicating the moving average. A red line on each plot indicates the optimum model according to multi objective optimization, as detailed in the methods. Plots are organized in rows for variable subsets and columns for ML algorithms.
